# Factors associated with country-variation in COVID-19 morbidity and mortality worldwide: an observational geographic study COVID-19 morbidity and mortality country-variation

**DOI:** 10.1101/2020.05.27.20114280

**Authors:** H Bellali, N Chtioui, M Chahed

**Author notes:** **Corresponding author:** Hedia Bellali, Epidemiology and Statistic Department, A Mami Hospital, Ariana 2080, Tunisia.

## Abstract

**Background:** The world is threatened by the outbreak of coronavirus disease 19 (COVID-19) since December 2019. The number of cases and deaths increased dramatically in some countries from March 2020. The objective of our study was to examine potential associated factors with country-variation in COVID-19 morbidity and mortality in the world.

**Methods:** We performed a retrospective geographic study including all countries with the most recent available data on free access on the web. We analyzed univariate and multivariable correlation between both the number of reported cases and deaths by country and demographic, socioeconomic characteristics, lockdown as major control measure, average annual temperature and relative humidity. We performed simple linear regression, independent t test and ANOVA test for univariate analyses and negative binomial regression model for multivariable analyses.

**Results:** We analyzed data of 186 countries from all world regions. As of 13^th^ April 2020, a total of 1 804 302 COVID-19 cases and 113 444 deaths were reported. The reported number of COVID-19 cases and deaths by countries was associated with the number of days between the first case and lockdown, the number of cases at lockdown, life expectancy at birth, average annual temperature and the socio-economic level. Countries which never implemented BCG vaccination reported higher mortality than others.

**Conclusions:** The pandemic is still ongoing and poses a global health threat as there is no effective antiviral treatment or vaccines. Thus, timing of control measure implementation is a crucial factor in determining the spread of the epidemic. It should be a lesson for this pandemic and for the future.

## 1. Introduction

The outbreak of a novel coronavirus (COVID-19) since the end of December has posed a significant threat to international health and the world economy. Almost the whole world is under lockdown for several weeks to contain the COVID-19 outbreak (1). As there is no vaccine, no effective specific treatment, the level of transmission reduction is crucial. Thus, interventions to mitigate the epidemic including social distancing by either localized or national lockdown were implemented by many countries (2).

Since the beginning of March, many countries have been hit particularly hard by COVID-19 including Europe and the United States of America. However, there are major country differences in both the spread of the infection and the mortality due to the coronavirus. Few univariable (3) and national (4,5) studies tried to explain the reasons of these observed differences. Socioeconomic, demography, health system and climate environment could be important in understanding the dramatically increased number of cases and deaths in some countries.

The aim of our study was to examine country-variation in COVID-19 morbidity and mortality in the world by analyzing 1) demographic, socioeconomic characteristics, 2) restrictions and time between the first case reported and restrictions decision 3) climate environment.

## 2. Methods

### 2.1. Study design

We performed a retrospective geographic study including all countries with the most recent available data on free access on the web. We analyzed univariate and multivariable correlation between both the number of cases and deaths by country and demographic, socioeconomic characteristics, lockdown as major control measure and climate environment.

### 2.2. Data collection

Data about all countries was collected from different sources. We downloaded the geographic distribution of cases and deaths worldwide data as of April, 13^th^ from the ECDC website (6). We aggregated data by country calculating the total reported cases and deaths for each country.

We collected population density, infant mortality rate 2015-2020 per 1000 inhabitant, the proportion of male/female, the proportion of people aged 65 years and over and the life expectancy at birth from the United Nations Website (7).

Human development index (HDI) for 2018-2019 was introduced into data from the report of the United Nations (8). We collected information on Universal BCG vaccination policy by countries from the BCG World Atlas (9).

We included the 2019 socioeconomic level classification for each country from the World Bank (10).

Information on lockdown was found on the BBC website (1) and was completed from other sources for the non-available information (11). We calculated the number of days between the first case and the lockdown and the total number of cases at lockdown from the above source and the geographic distribution of cases data which includes the daily number of cases and deaths by country.

We collected average yearly temperature in °C for 2019 for all countries from (12) and 2019 average annual relative humidity [2 m] in percentage from (13).

### 2.3. Data analyses

The dependent variables were the number of cases and the number of deaths by country. The independent variables by country were the population size, the population density, the infant mortality rate 2015-2020, the percentage of male/female, the percentage of people aged 65 years and over, the life expectancy at birth, human development index, socioeconomic classification (low income, middle income, high income), Universal BCG vaccination policy (yes/no), lockdown (none, localized or national lockdown), number of days between the first case and lockdown and the number of cases at lockdown.

We performed simple linear regression with exponential function, independent t test and ANOVA test for univariate analyses. We used generalized linear models with negative binomial distribution for multivariable analyses. We included into the model all significant associated variables from the univariate analyses, we introduced interaction terms and we adjusted for the population size. The significant level of all tests was set at p value ≤ 0.05. We analyzed all data with SPSS Software.

## 3. Results

We analyzed data of 186 countries from all world regions. As of 13^th^ April 2020, a total of 1 804 302 COVID-19 cases and 113 444 deaths were reported. The United States of America reported the highest number of cases and deaths (557 571 cases and 22 108 deaths), Spain reported the second highest number of cases (166 019) and Italy reported the second highest number of deaths (19 901) (figures 1 & 2).

**Figure 1:**
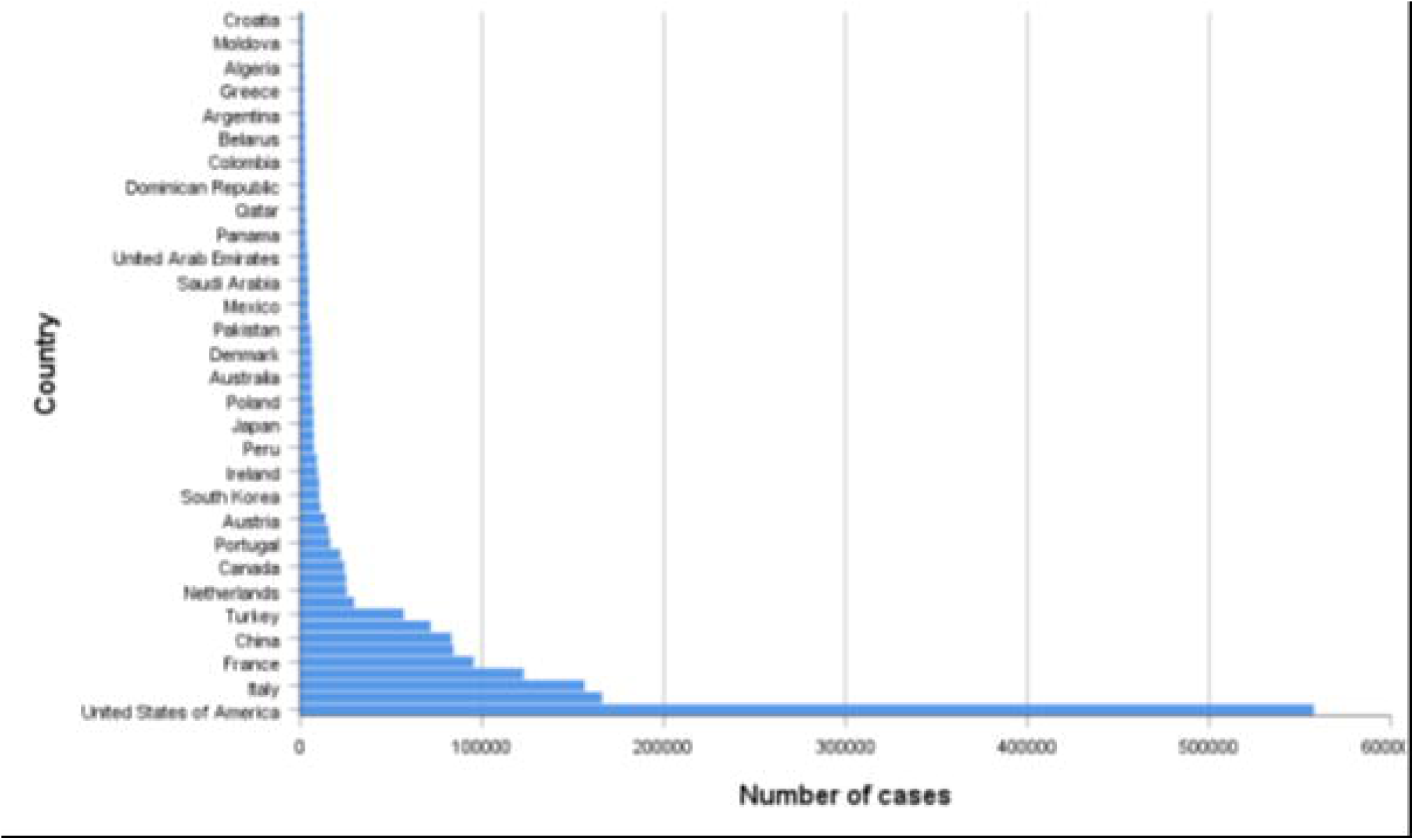
distribution of the number of COVID-19 cases by country, as of April 13^th^ 2020

**Figure 2:**
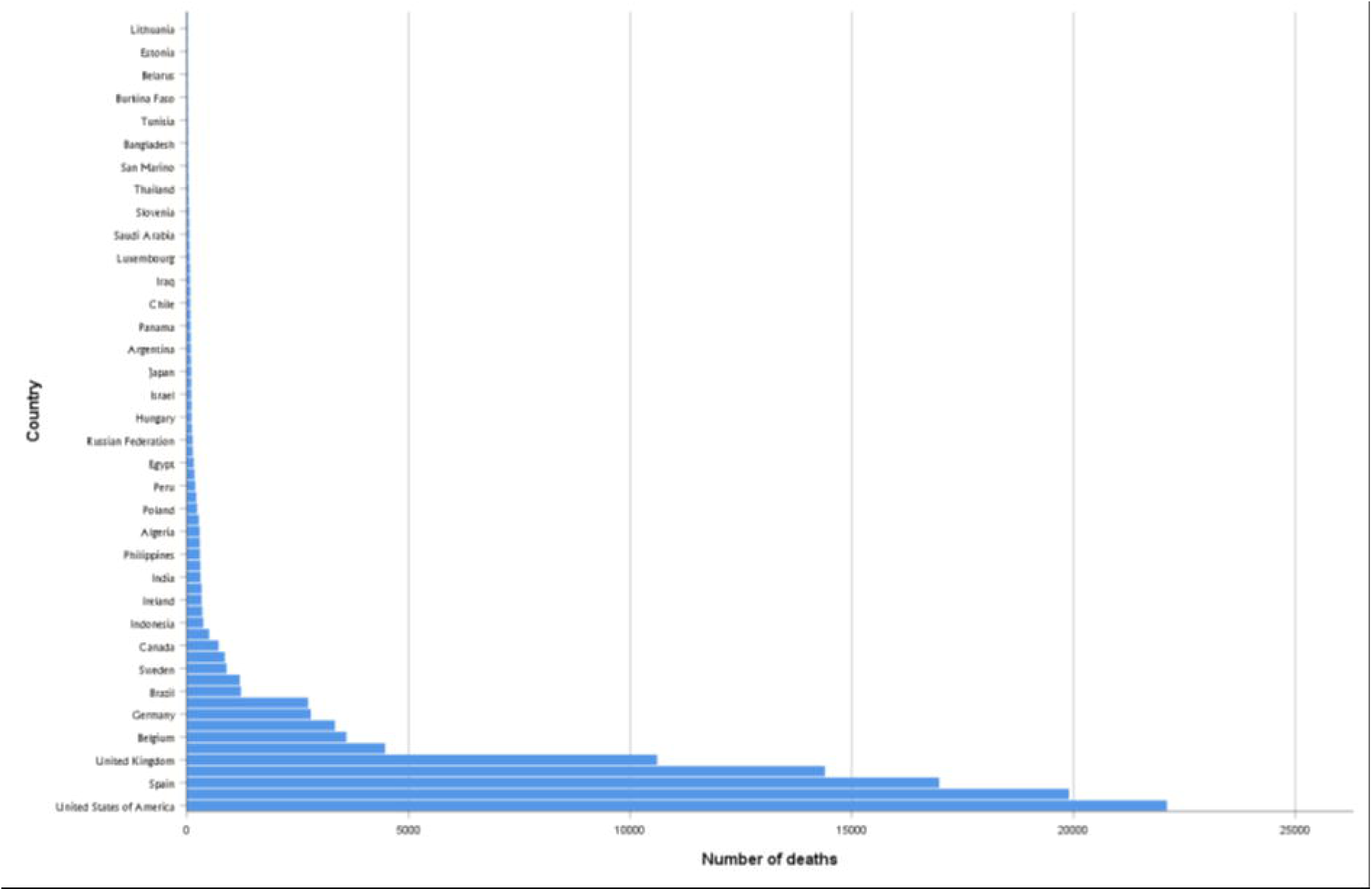
distribution of the number of COVID-19 deaths by country, as of April 13^th^ 2020

### Morbidity (Number of cases by country)

The number of COVOD-19 cases in this first univariate analysis was positively correlated to the number of cases at lockdown and the number of days between the first case and lockdown; correlation coefficient r=0.627, p value<10^-3^ and r=0.562, p value<10^-3^ respectively. The number of reported cases was higher for countries with the highest number of reported cases at lockdown (figure 3) and the large time between the reported first case and lockdown (figure 4).

**Figure 3:**
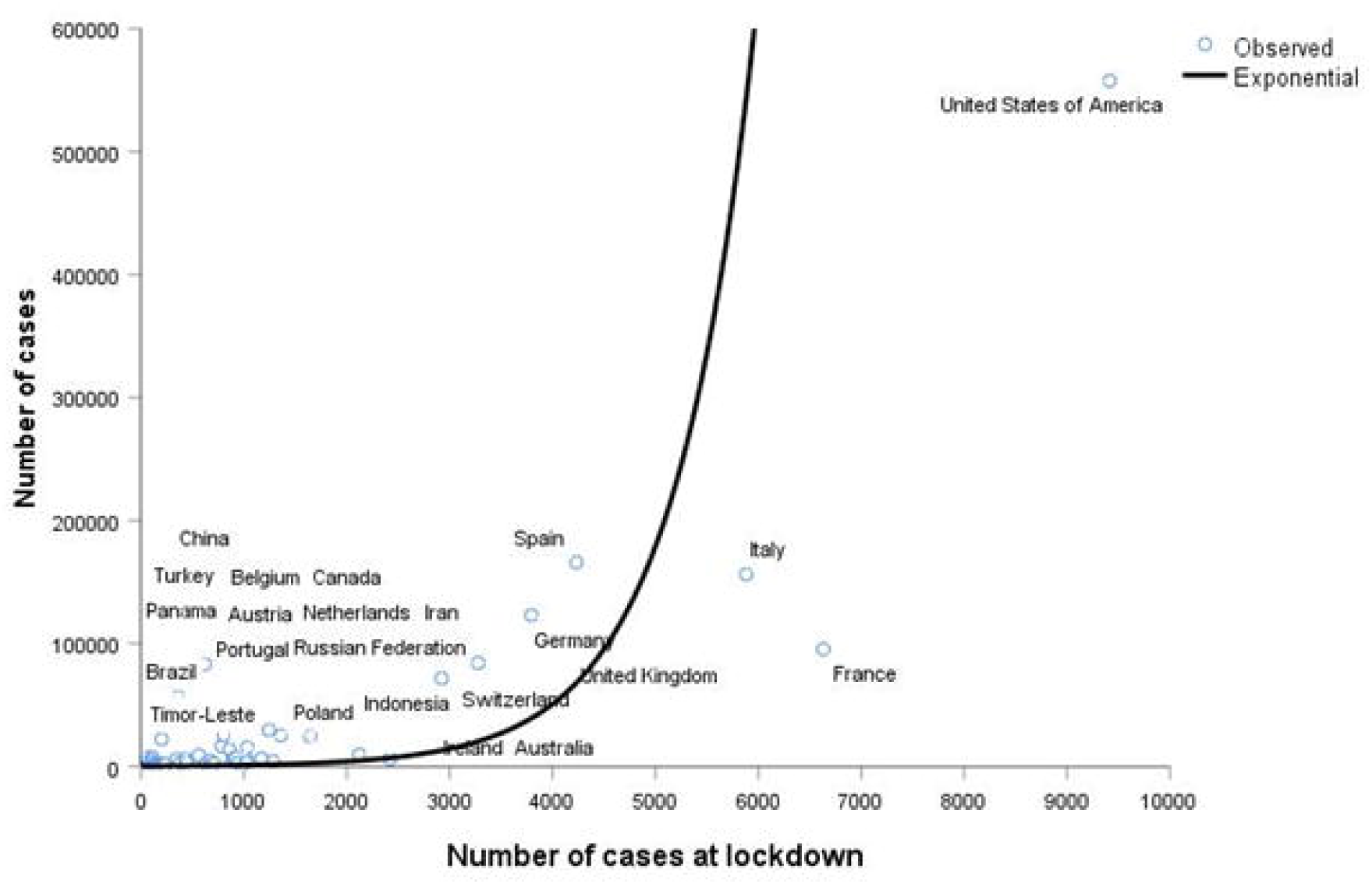
Correlation between the number of COVID-19 cases and the number of cases at lockdown

**Figure 4:**
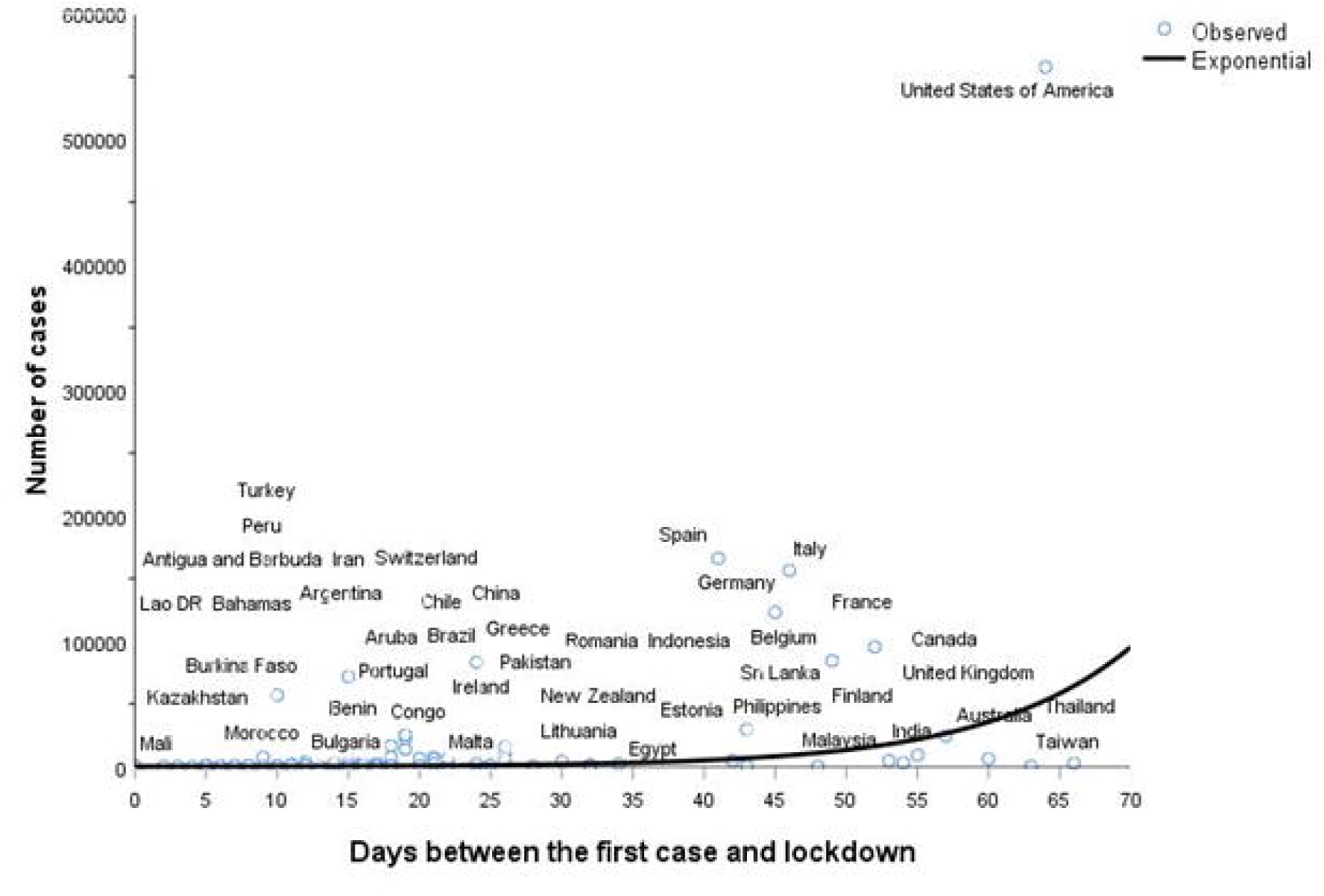
Correlation between the number of COVID-19 cases and the number of days between the first case and lockdown

Infant mortality was negatively correlated with the number of COVID-19 cases (r=-0.548, p value<10^-3^). The number of COVID-19 infection was higher among countries with low infant mortality rate (figure 5).

**Figure 5:**
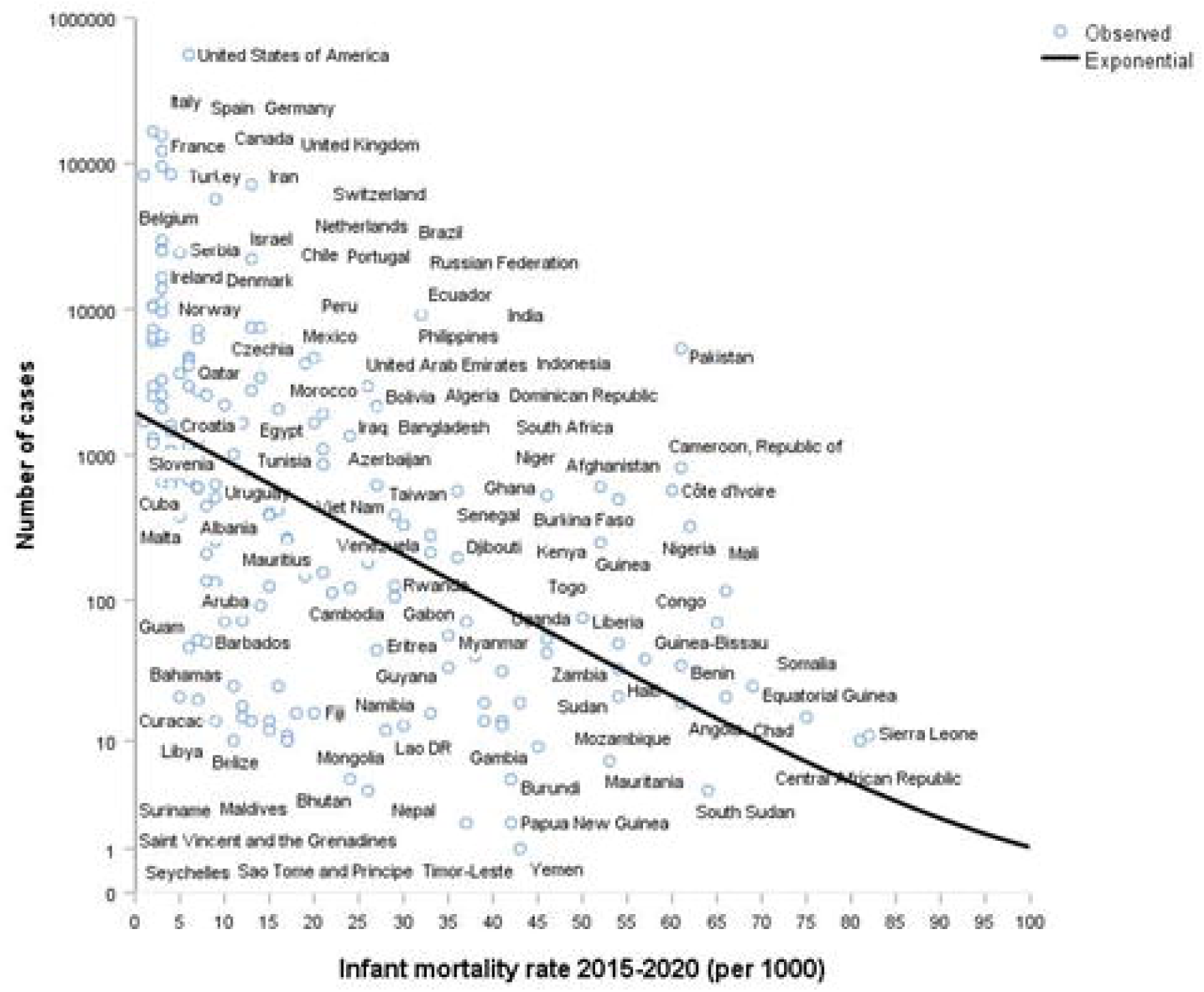
Correlation between the number of COVID-19 cases and the infant mortality rate We also observed positive correlation with HDI (r=0.670, p<10^-3^), life expectancy at birth (r=0.600, p<10^-3^) and percentage of people aged 65 years and over (r=0.578, p<10^-3^) (figures 6, 7 & 8).

**Figure 6:**
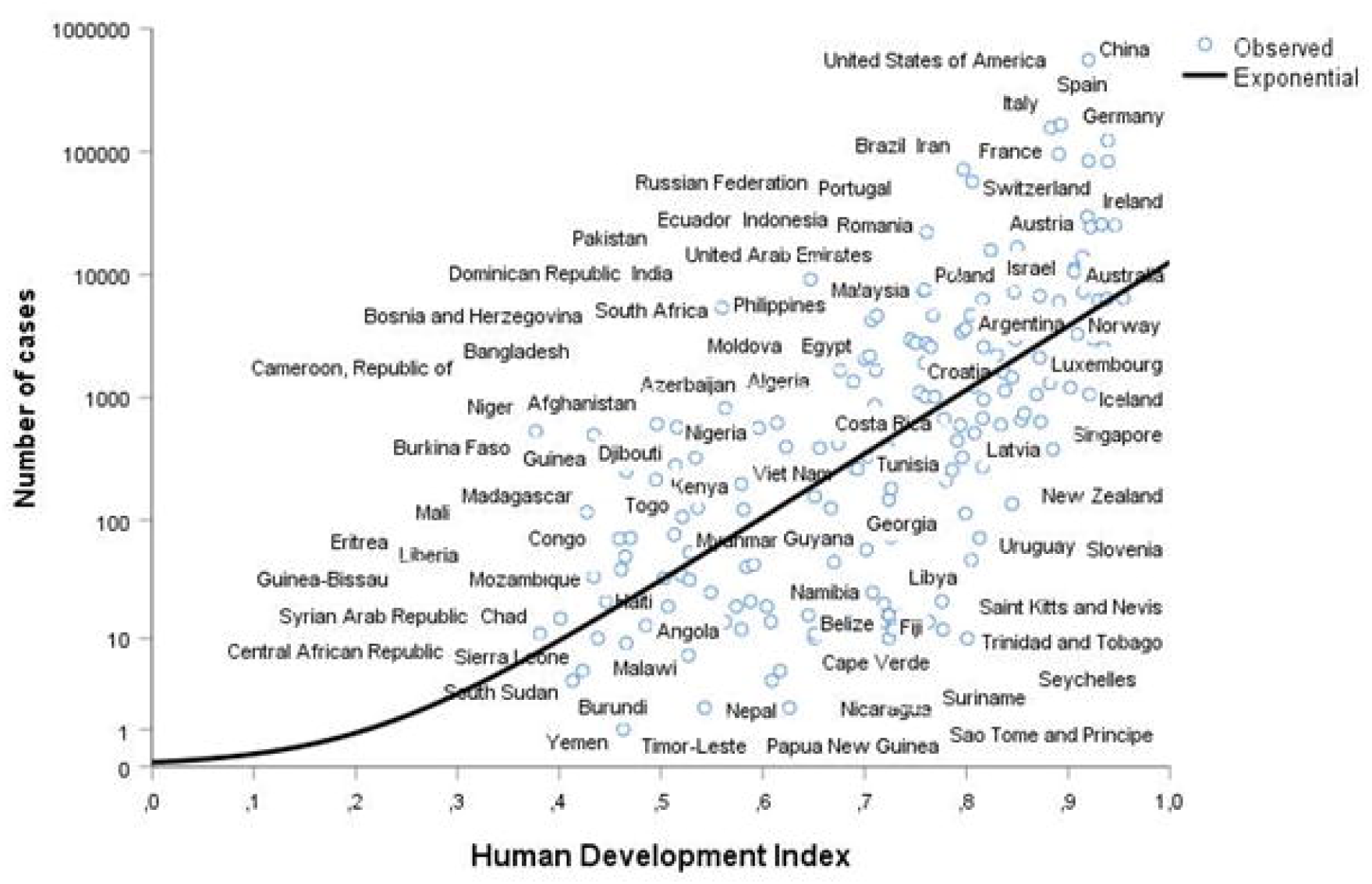
Correlation between the number of COVID-19 cases and the HDI

**Figure 7:**
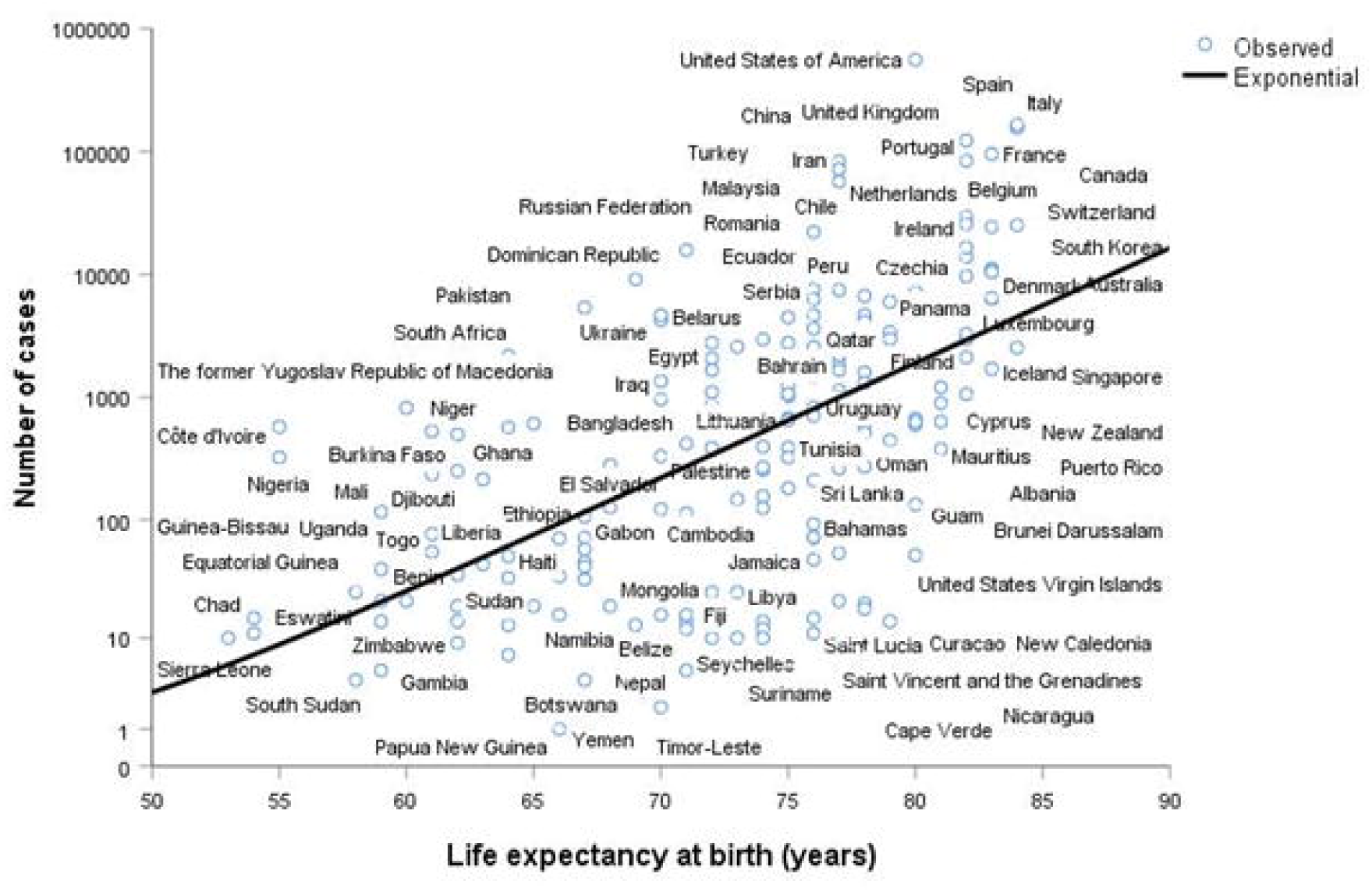
Correlation between the number of COVID-19 cases and the life expectancy at birth

**Figure 8:**
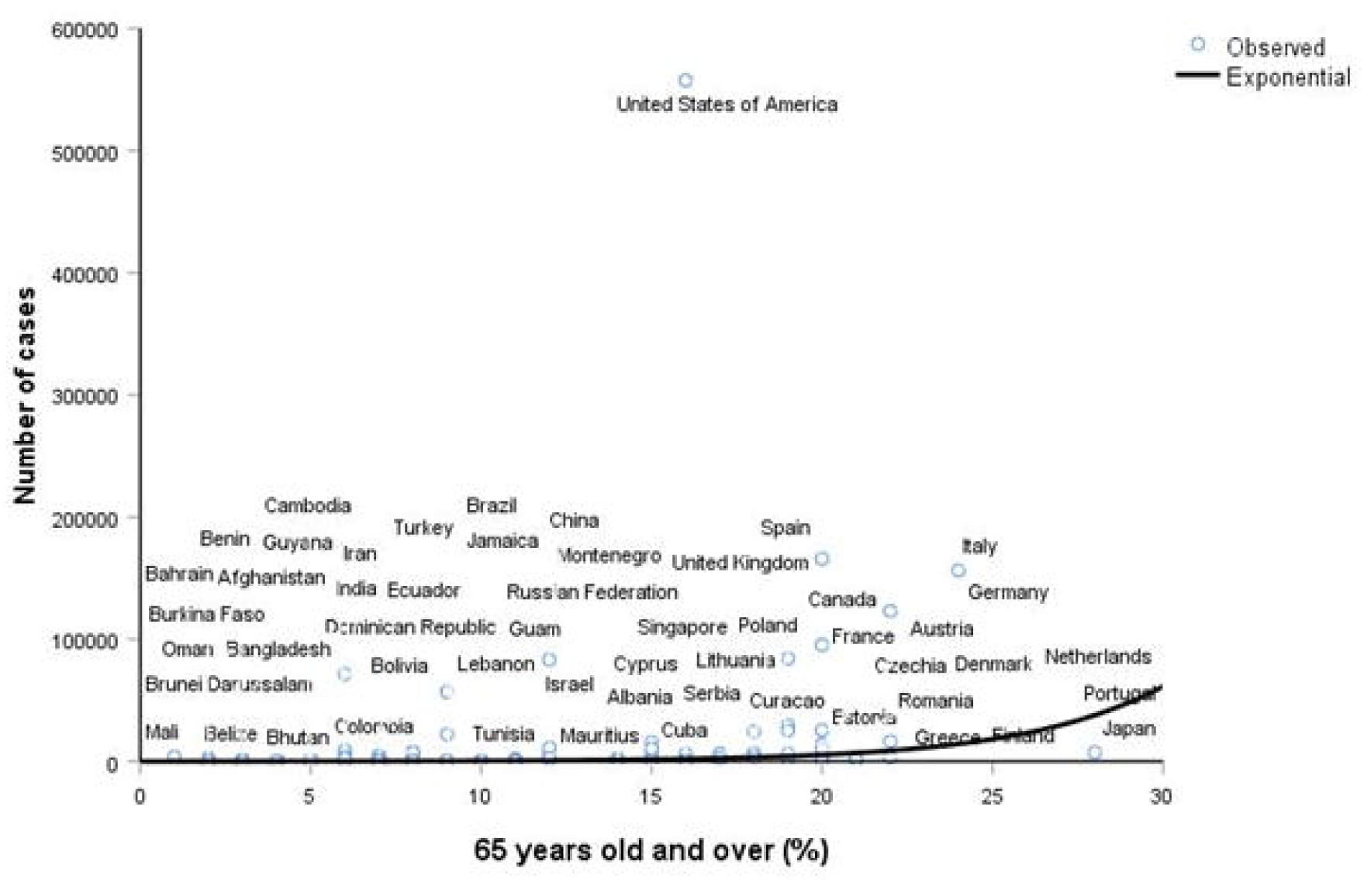
Correlation between the number of COVID-19 cases and the proportion of people aged 65 years and over

A negative correlation was observed between the average annual temperature and the number of COVID-19 cases (r=-0.504, p<10^-3^). The number of reported COVID-19 cases was higher in countries with low average yearly temperature (figure 9).

**Figure 9:**
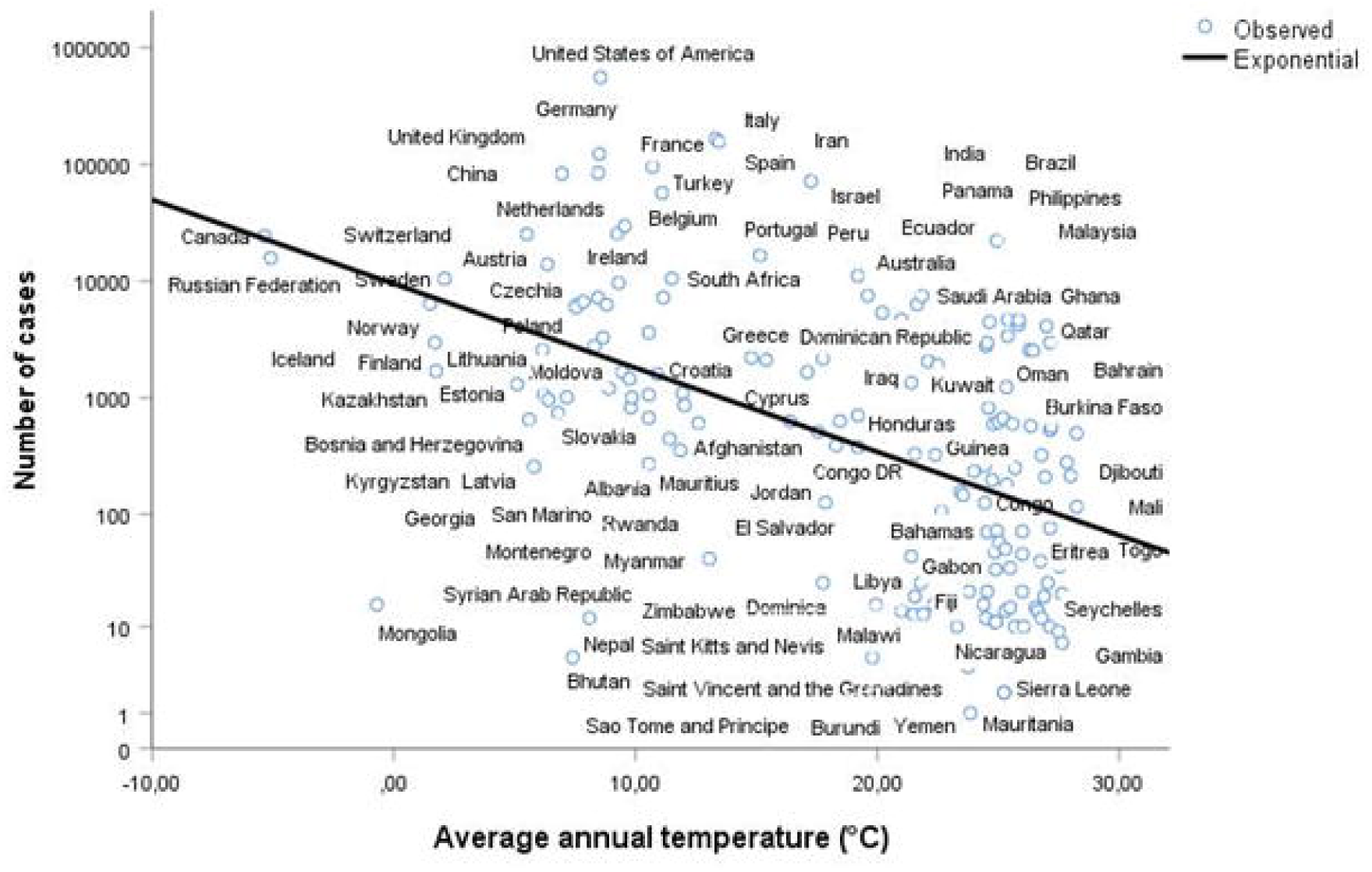
Correlation between the number of COVID-19 cases and the average annual temperature

The average number of cases in high income countries was 23 001.40, 3 782.65 in middle income countries and 114.27 in low income countries, the difference was significant p value=0.018. The average number of reported cases in countries with and without universal BCG vaccination policy was respectively 2930.70 and 69 203.95; p value=0.04 for the global population. After stratification by socioeconomic level, the difference was significant only for high income countries; p value=0.041.

There was no significant correlation between the number of cases and lockdown, population density, average annual relative humidity and percentage of male/female.

The final negative binomial regression model, indicated that the number of days between the first case and lockdown, the number of cases at lockdown, life expectancy at birth, average annual temperature and the economic level were independently associated with the reported number of COVID-19 cases by countries (table 1).

**Table 1:** Factors associated with country-variation in COVID-19 morbidity

| Factor | RR<sub>a</sub>* | 95% CI** | p value |
| --- | --- | --- | --- |
| Number of cases at lockdown | 1.001 | 1.000-1.001 | <10<sup>-3</sup> |
| Number of days between the first case and lockdown | 1.031 | 1.012-1.049 | 0.001 |
| Life expectancy at birth | 1.158 | 1.122-1.196 | <10<sup>-3</sup> |
| Socioeconomic level | 0.189 | 0.114-0.313 | <10<sup>-3</sup> |
| Average annual temperature | 0.928 | 0.901-0.956 | <10<sup>-3</sup> |

\*Adjusted Risk Ratio; \*\*95% Confidence Interval

### Mortality (Number of deaths by country)

Univariate analysis showed that the number of COVOD-19 deaths was positively correlated to the number of cases at lockdown and the number of days between the first case and lockdown; correlation coefficient r=0.906, p value<10^-3^ and r=0.445, p value<10^-3^ respectively. The number of reported deaths was higher for countries with the highest number of reported cases at lockdown (figure 10) and the large time between the reported first case and lockdown (figure 11).

**Figure 10:**
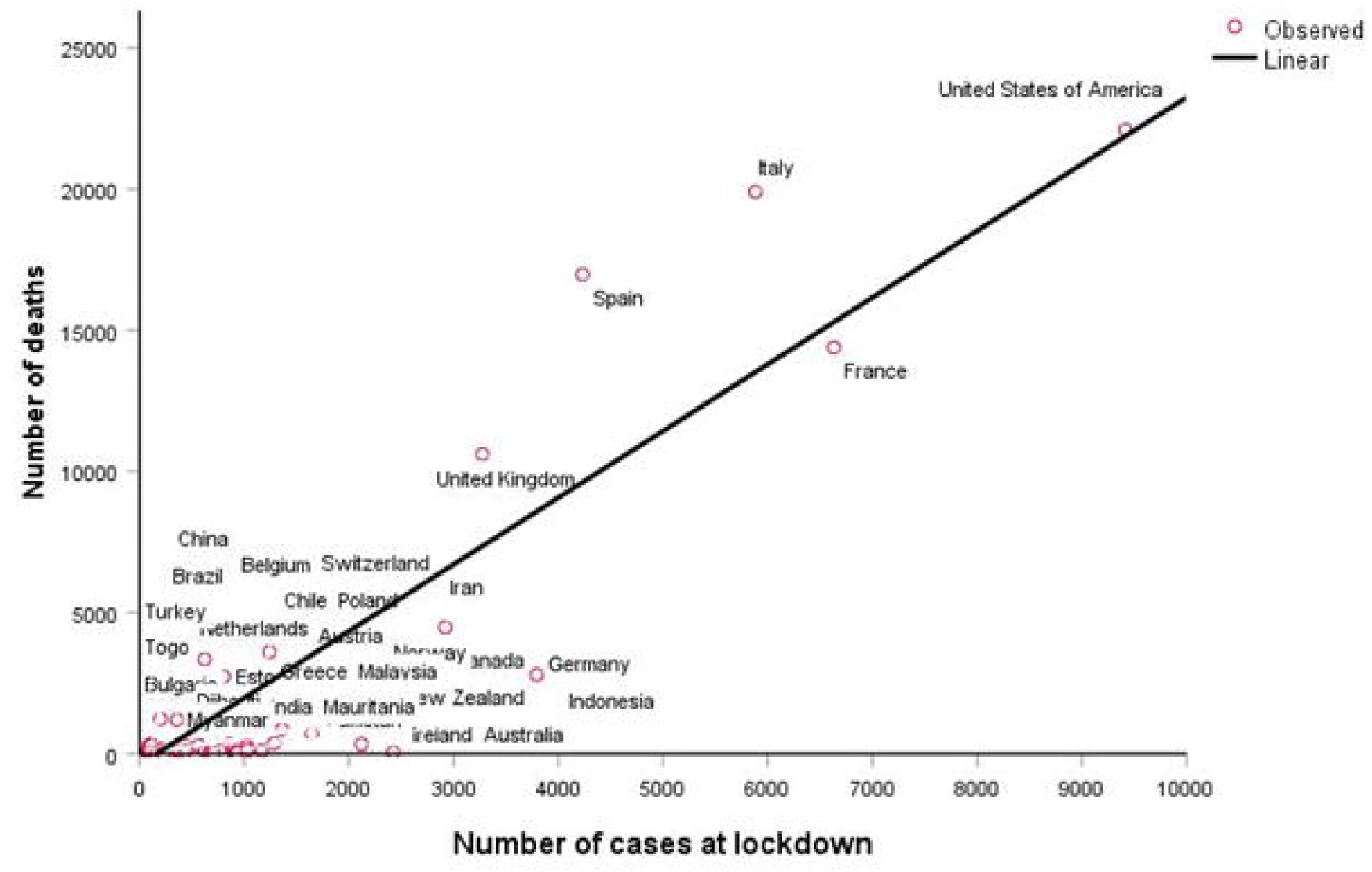
Correlation between the number of COVID-19 deaths and the number of cases at lockdown

**Figure 11:**
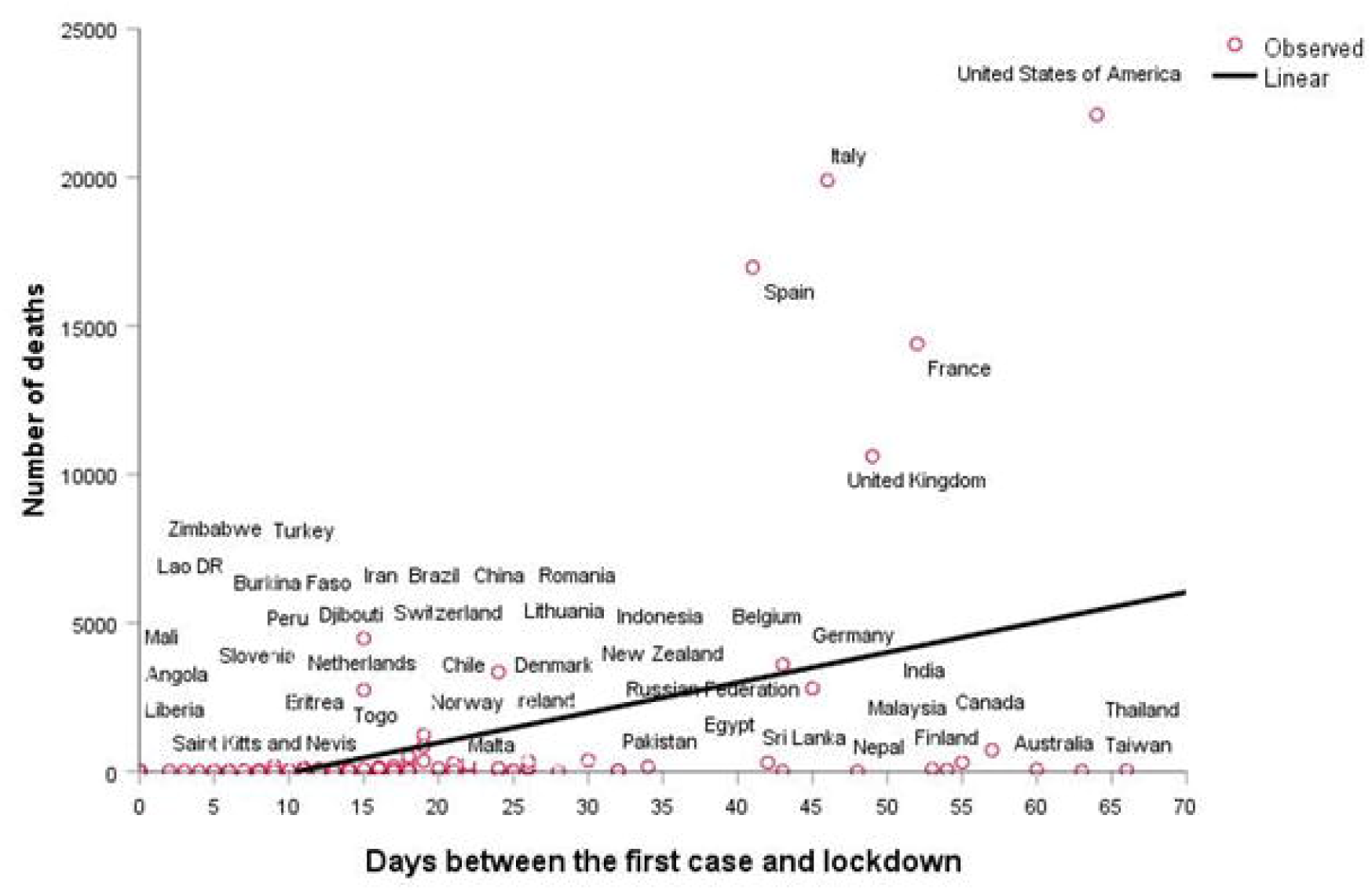
Correlation between the number of COVID-19 deaths and the number of days between the first case and lockdown

Infant mortality was negatively correlated with the number of COVID-19 deaths (r=-0.184, p value=0.014). The number of COVID-19 deaths was higher among countries with low infant mortality rate (figure 12).

**Figure 12:**
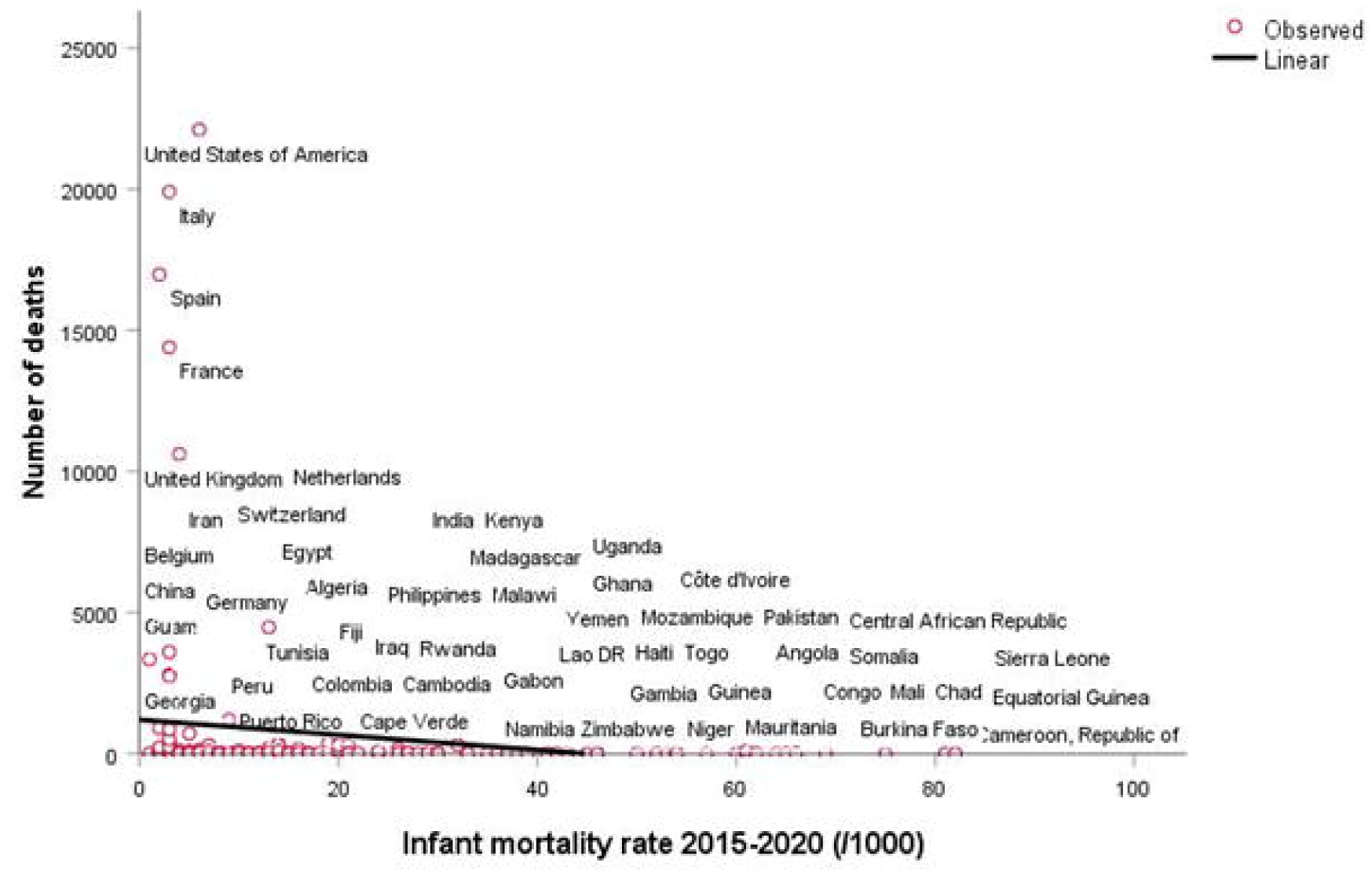
Correlation between the number of COVID-19 deaths and the infant mortality rate We also observed positive correlation with HDI (r=0.257, p=0.001), life expectancy at birth (r=0.257, p<10^-3^) and percentage of people aged 65 years and over (r=0.308, p<10^-3^) (figures 13, 14 & 15).

**Figure 13:**
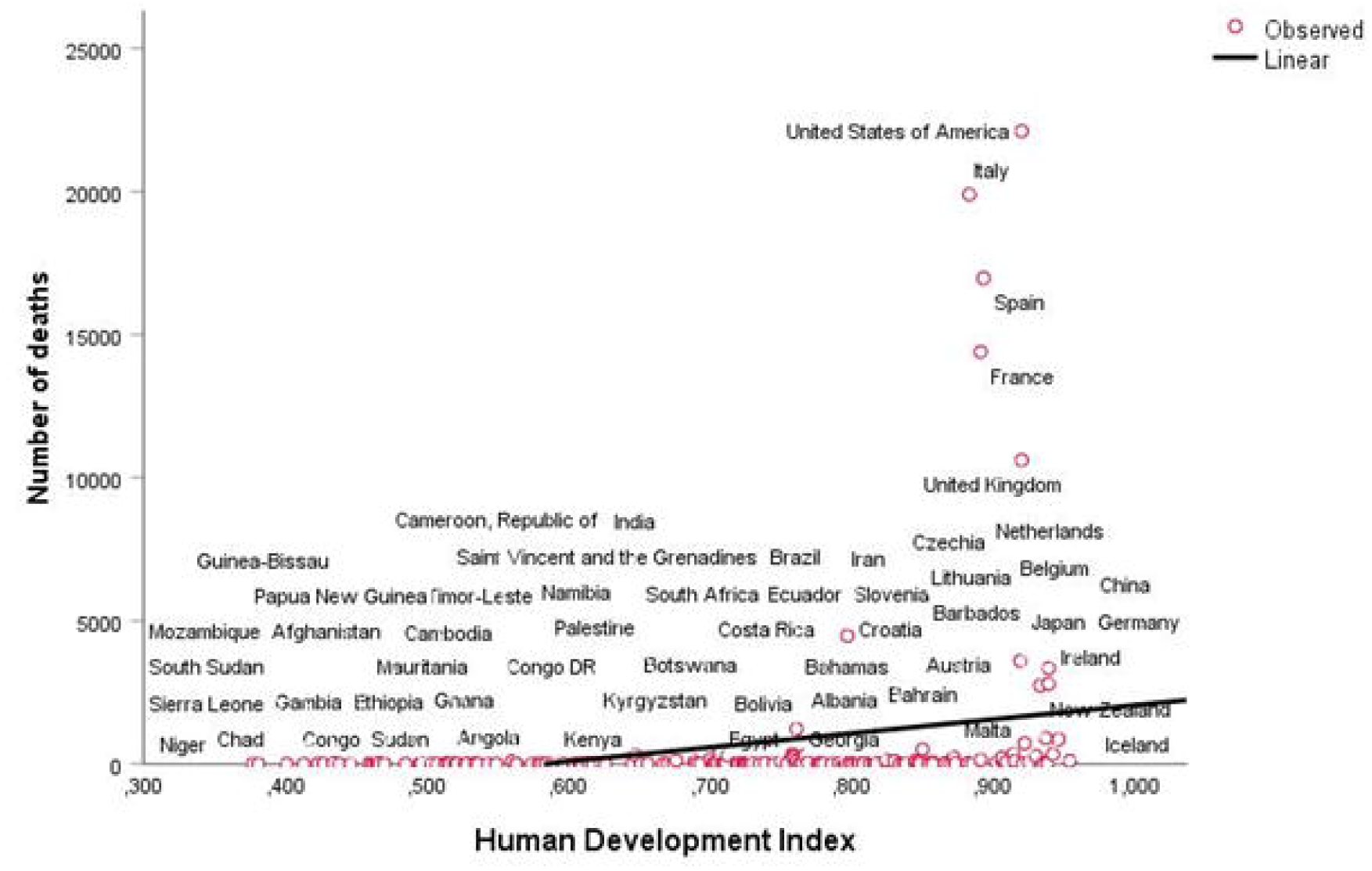
Correlation between the number of COVID-19 deaths and the HDI

**Figure 14:**
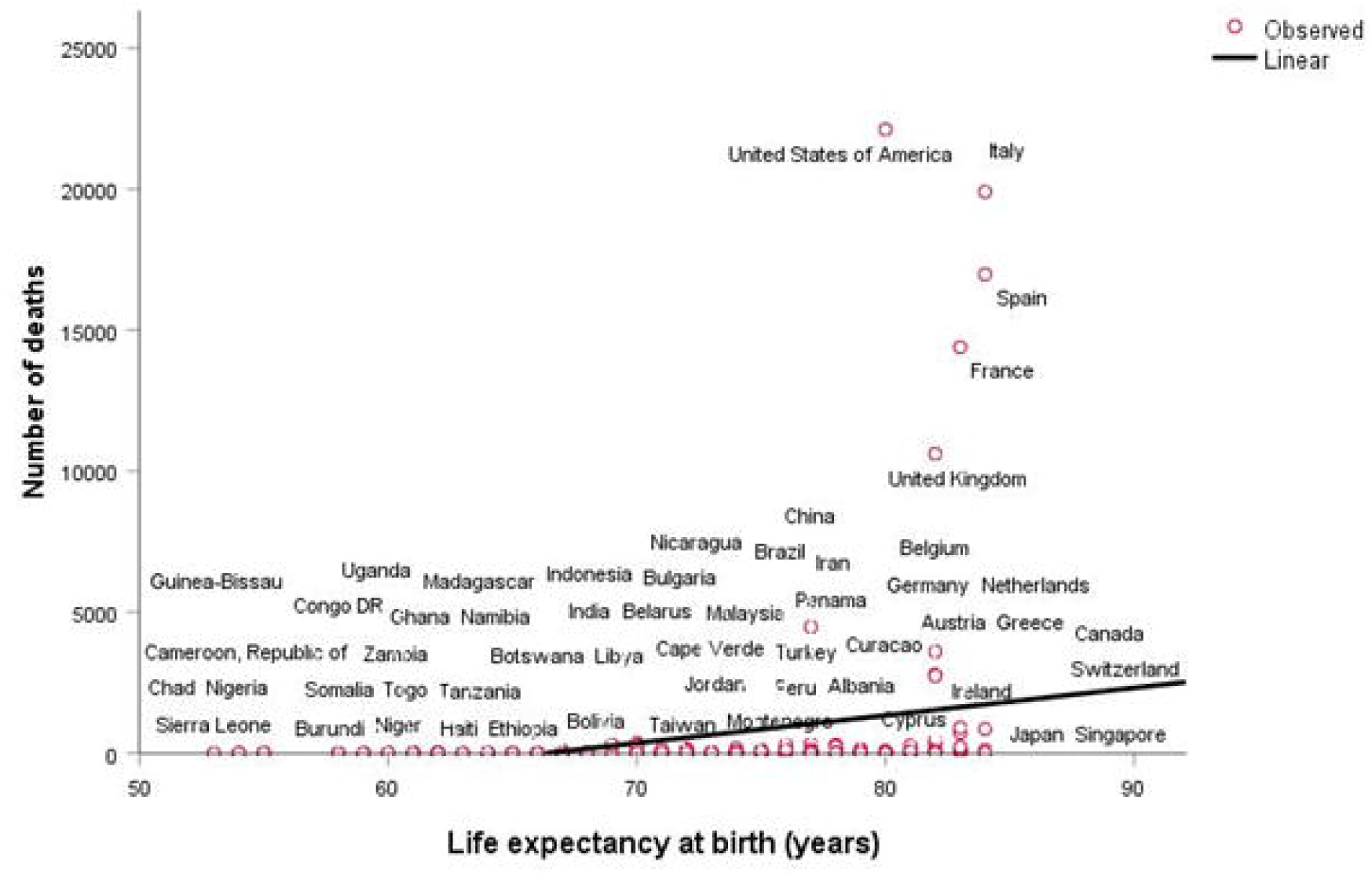
Correlation between the number of COVID-19 deaths and the life expectancy at birth

**Figure 15:**
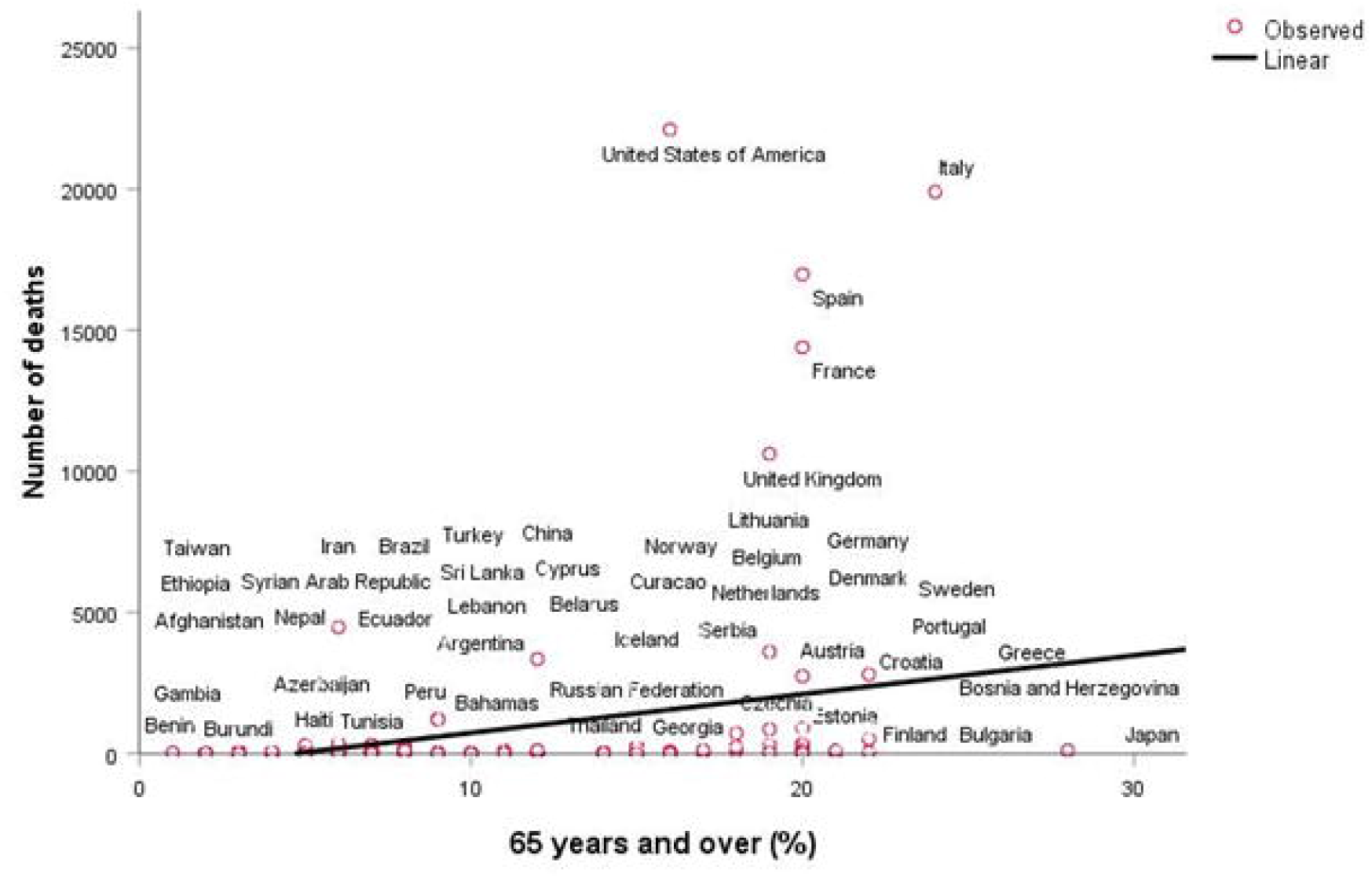
Correlation between the number of COVID-19 deaths and the proportion of people aged 65 years and over

A negative correlation was observed between the average annual temperature and the number of COVID-19 deaths (r=-0.199, p=0.199). The number of reported COVID-19 deaths was higher in countries with low average yearly temperature (figure 16).

**Figure 16:**
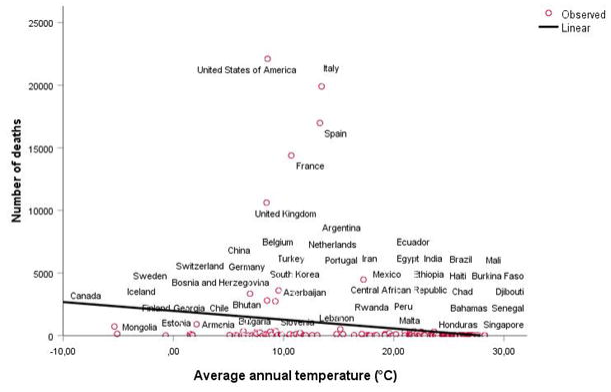
Correlation between the number of COVID-19 deaths and the average annual temperature

The average number of deaths in high income countries was 1 569.95, 155.5 in middle income countries and 3.9 in low income countries, the difference was significant p value=0.004. The average number of reported deaths in countries with and without universal BCG vaccination policy was respectively 104.31 and 5 053.89; p value=0.011 for the global population. After stratification by socioeconomic level, the difference was significant only for high income countries; p value=0.010.

There was no significant correlation between the number of deaths and lockdown, population density, average annual relative humidity and percentage of male/female.

Multivariable analyses using negative binomial regression model found that the number of days between the first case and lockdown, the number of cases at lockdown, life expectancy at birth, average annual temperature, the economic level and the universal BCG vaccination policy were independently associated with the reported number of COVID-19 deaths by countries (table 2).

**Table 2:**
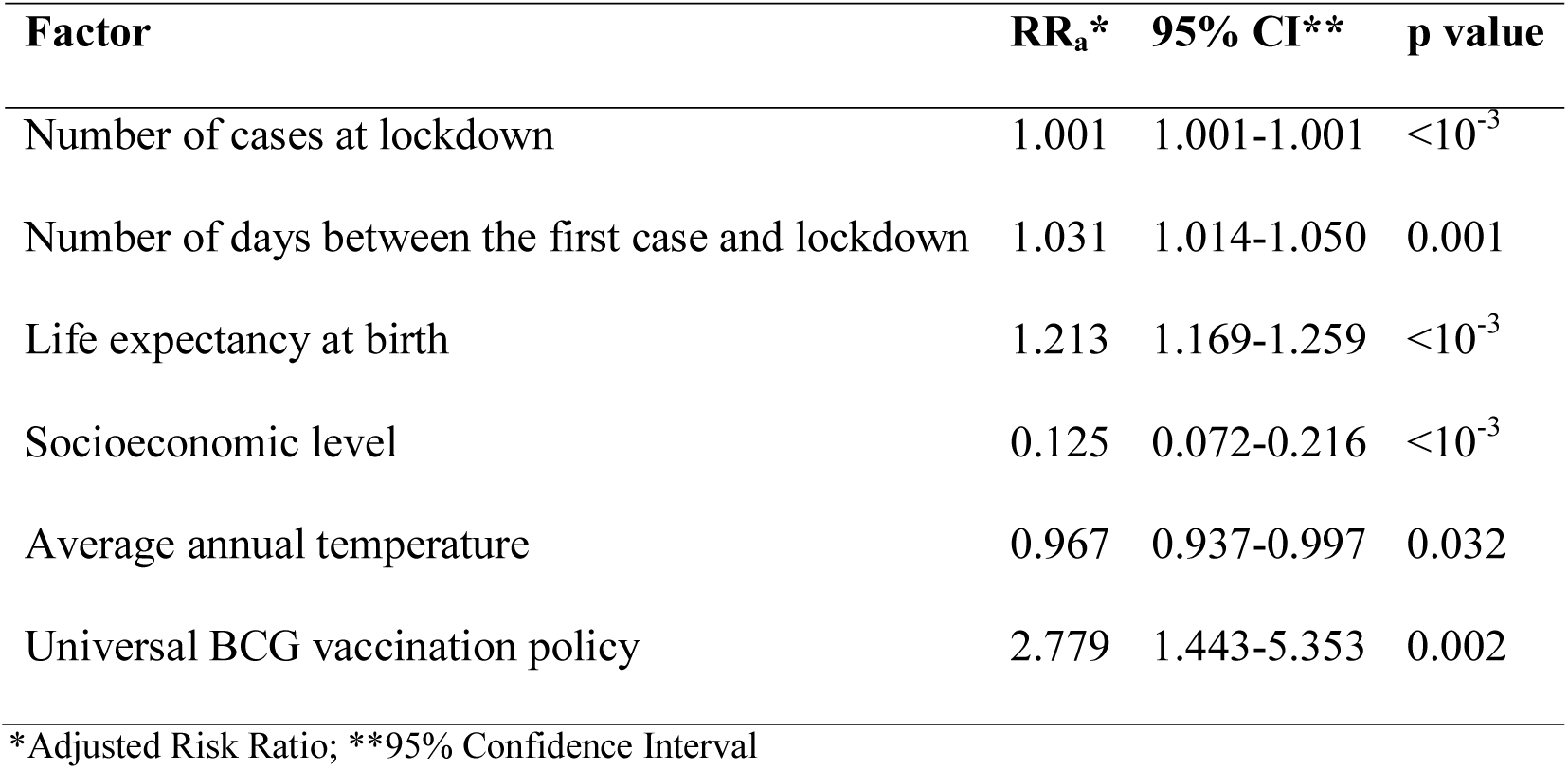
Factors associated with country-variation in COVID-19 mortality

| Factor | RR<sub>a</sub>* | 95% CI** | p value |
| --- | --- | --- | --- |
| Number of cases at lockdown | 1.001 | 1.001-1.001 | <10<sup>-3</sup> |
| Number of days between the first case and lockdown | 1.031 | 1.014-1.050 | 0.001 |
| Life expectancy at birth | 1.213 | 1.169-1.259 | <10<sup>-3</sup> |
| Socioeconomic level | 0.125 | 0.072-0.216 | <10<sup>-3</sup> |
| Average annual temperature | 0.967 | 0.937-0.997 | 0.032 |
| Universal BCG vaccination policy | 2.779 | 1.443-5.353 | 0.002 |

\*Adjusted Risk Ratio; \*\*95% Confidence Interval

## 4. Discussion

In this study we investigated the potential factors that can explain the country-variation in COVID-19 reported cases and deaths worldwide. We included the maximum of variables which are already known to make differences between countries in terms of infectious morbidity and mortality; such as the performance of the health system, population sociodemographic characteristics, socioeconomic level and climate variability. Infant mortality rate and life expectancy at birth are related to the performance of country health system (14). Population sociodemographic characteristics include human development index, the proportion of male/female, the proportion of people aged 65 years and over and the population density and size. These indicators were used mainly to adjust for confounders. Temperature and relative humidity are already known as important factors of survival and transmission of SRAS and MERS coronaviruses (15–19). Control measures and their timeline of application may also be different between countries; we explored the lockdown and the time of its implementation regarding the ongoing epidemic as a major intervention. We included the universal BCG vaccination policy to assess its effect on the number of cases and deaths.

Our study showed that the differences between countries in COVID-19 transmission and mortality could be partially explained by the early decision of lockdown, life expectancy at birth, average annual temperature and the economic level. The universal BCG vaccination policy was only associated with deaths; countries which never implemented BCG vaccination reported higher mortality than others.

We observed that countries that implemented BCG vaccination in their national immunization schedule reported less deaths due to COVID-19. This finding suggests that BCG vaccination doesn’t protect against COVID-19 infection susceptibility but might decrease the severity and consequently reducing mortality by COVID-19. A study (20) carried out in Guinea-Bissau concluded that BCG vaccine has protective effect on acute lower respiratory tract infection. Another research (21) conducted in Spain concluded that BCG vaccination at birth may decrease respiratory infection and sepsis hospitalization. Similarly, Niobey et al (22) found an association between BCG immunization and a reduced risk of pneumonia mortality in children in Brazil. The mechanism of this effect is not clearly explained, but it seems to be related to the reinforcement of the nonspecific immune response (23).

The situation of lockdown was not associated with the morbidity and the mortality by the COVID-19. In fact, the majority of countries took the decision to shut down all their activities in time of the ongoing epidemic. However, this study concluded that the time of the nationwide lockdown and consequently, the number of cases reported in the country when the lockdown was implemented, were strongly associated with COVID-19 mortality and morbidity. The high number of cases at lockdown and the large number of days between the first case reported in the country and the lockdown results in more reported cases and deaths. This finding is rational, an epidemic follow the natural growth pattern: the more infected, the more infections. The Basic Reproduction Number R0 is the expected number of secondary cases generated by one infected subject. It depends on the transmissibility, the average rate of contact between susceptible and infected individuals, and the duration of infectiousness (24,25).

It suggests that the early decision of maximum containment measures (Stay at home recommendations) decrease the transmission in the community, reduce the burden of the disease to hospitals and improve the prognosis resulting in less mortality (26). Early implementation of control measures assume that the transmission can occur from asymptomatic COVID-19 carriers to the community since asymptomatic COVID-19 infection is possible (27,28) or before and at the beginning of symptoms onset (29).

Tian et al (30) found that control measures were strongly associated with the containment of COVID-19 avoiding hundreds of thousands incident cases.

Carlo Signorelli et al (31) confirmed that the timing of mitigation measures is of high importance in reducing the transmission in Italy. The containment measures are most effective if they are decided and implemented at an early stage of the epidemic and on large areas. Another study in Italy (32) indicated that a stronger effect on the epidemic growth was obtained when the lockdown was enforced earlier. In Spain and Italy (33), implementation of more restrictive measures resulted in a change in the trend slopes in daily incident cases and intensive care unit admissions. Several works (34–39) concluded that the effect of community mitigation measures and potential impacts on community mobility, depends greatly on the choice of date, early implementation of the most restrictive measures may be more effective. It has a strong potential to reduce the magnitude of the epidemic which is particularly important, as this reduces the acute pressure on the health-care system and improve the prognosis of severe cases.

Our work indicated that the average annual temperature is negatively associated with the COVID-19 morbidity and mortality. It suggests that a difference of 1°C in temperature, decrease the number of reported COVID-19 cases and deaths of 7.2% and 3.3% respectively. The effect of average temperature on the COVID-19 morbidity and mortality was consistent with that found in other country level studies (40,41). The differences with our results can be related to the use of different outcome measures and methods and to the number of variables included into the analysis, we adjusted for more variables that are basically known to have relationship mainly with death and which vary from one country to another.

The negative association of average temperature with COVID-9 infection was observed in many other studies (4,42–47). It seems that high temperature shortens the period of survival in the environment of the coronavirus resulting in the decrease of the potential transmissibility. Our study indicated that countries from the group “high income” had less COVID-19 cases and death comparing with middle/low income countries. This finding can be explained by the differences in both educational level and lifestyle factors (48,49).

We observed that the indicator of « life expectancy at birth » was positively associated with COVID-19 mortality more than morbidity. An increase of one year raises the number of death by 21.3% and the number of cases by 15.8%. This result suggests that if people are expected to live for a long time, they will be more susceptible to develop severe infections and to die.

Our work had several limitations. First, we carried out an observational geographic study. Our results cannot prove causality, they just suggest an association. However, in such pandemic situation due to emerging pathogen, these results are useful because data are immediately available and could help predicting the spread of the epidemic and guide control measures in the absence of effective treatment and vaccine to tackle the epidemic. Second, we used general indicators by countries such as average annual temperature and relative humidity; the data could be not very precise. But it might not affect the analysis since the objective was to compare outcomes between countries once we used the same variable and the same source for all countries. Third, we didn’t take into account the difference in population rate change in behaviors (50), societal and social psychological factors (51) and the real application and respect for total containment, that’s why our work explained only a part of country-variation on COVID-19 morbidity and mortality.

## 5. Conclusions

Despite these limitations, our study highlights the importance of early and efficient implementation of control measures to curb the epidemic by reducing the transmission, new incident cases and deaths. If the BCG vaccine can boost the non-specific immune response, it can be used to lessen the severity of COVID-19 infection, particularly in high risk groups in order to decrease the number of severe cases, hospitalization and death. Association between high temperature and the COVID-19 morbidity and mortality should be interpreted with caution. At least, it can help authorities to decide for when and how to lift lockdown, mainly for developing countries with socio-economic difficulties.

## Data Availability

All data referred to in the manuscript are available and could be uploaded in excel sheet.

## Declaration of competing interest

The authors declare that they have no competing interests related to this work.

## Funding

This work didn’t need any funding source.

## Notes

### Competing Interest Statement

The authors have declared no competing interest.

### Funding Statement

No external funding was received for this work.

### Author Declarations

We didn't use human informations nor samples. We used geographic cumulative cases and deaths distribution from the ECDC website and we informed them and got the permission for publication.

## References

1. The world in lockdown in maps and charts. BBC News [Internet]. 2020 Apr 7 [cited 2020 Apr 21]; Available from: https://www.bbc.com/news/world-52103747

2. Ferguson N, Laydon D, Nedjati Gilani G, Imai N, Ainslie K, Baguelin M, et al. Report 9: Impact of non-pharmaceutical interventions (NPIs) to reduce COVID19 mortality and healthcare demand [Internet]. Imperial College London; 2020 Mar [cited 2020 Apr 21]. Available from: http://spiral.imperial.ac.uk/handle/10044/1/77482

3. Miller A, Reandelar MJ, Fasciglione K, Roumenova V, Li Y, Otazu GH. Correlation between universal BCG vaccination policy and reduced morbidity and mortality for COVID-19: an epidemiological study [Internet]. Epidemiology; 2020 Mar [cited 2020 Apr 21]. Available from: http://medrxiv.org/lookup/doi/10.1101/2020.03.24.20042937

4. Shi P, Dong Y, Yan H, Zhao C, Li X, Liu W, et al. Impact of temperature on the dynamics of the COVID-19 outbreak in China. Sci Total Environ [Internet]. 2020 Aug 1 [cited 2020 Apr 28];728:138890. Available from: http://www.sciencedirect.com/science/article/pii/S0048969720324074

5. XieJ, Zhu Y. Association between ambient temperature and COVID-19 infection in 122 cities from China. Sci Total Environ [Internet]. 2020 Jul 1 [cited 2020 May 4];724:138201. Available from: http://www.sciencedirect.com/science/article/pii/S0048969720317149

6. Download today’s data on the geographic distribution of COVID-19 cases worldwide [Internet]. European Centre for Disease Prevention and Control. 2020 [cited 2020 Apr 21]. Available from: https://www.ecdc.europa.eu/en/publications-data/download-todays-data-geographic-distribution-covid-19-cases-worldwide

7. World Population Prospects - Population Division - United Nations [Internet], [cited 2020 Apr 21]. Available from: https://population.un.org/wpp/Download/Standard/Mortality/

8. 2018_human_development_statistical_update_fr.pdf [Internet]. Available from: http://hdr.undp.org/sites/default/files/2018_human_development_statistical_update_fr.pdf

9. BCG World Atlas [Internet], [cited 2020 Apr 21]. Available from: http://www.bcgatlas.org/index.php

10. World Bank Country and Lending Groups – World Bank Data Help Desk [Internet]. Available from: https://datahelpdesk.worldbank.org/knowledgebase/articles/906519-world-bank-country-and-lending-groups

11. Güner R, Hasanoǧlu I, Aktaş F. COVID-19: Prevention and control measures in community. Turk J Med Sei. 2020 21;50(SI-1):571–7.

12. Average Yearly Temperature by Country [Internet], [cited 2020 Apr 21]. Available from: https://statpedia.com/stat/Average_Yearly_Temperature_by_Country/HJd70cGK

13. ERA5 download Basel [Internet], meteoblue. [cited 2020 May 4], Available from: https://www.meteoblue.com/en/weather/archive/era5/basel_switzerland_2661604

14. Braithwaite J, Hibbert P, Blakely B, Plumb J, Hannaford N, Long JC, et al. Health system frameworks and performance indicators in eight countries: A comparative international analysis. SAGE Open Med [Internet]. 2017 Jan 4 [cited 2020 May 4];5. Available from: https://www.ncbi.nlm.nih.gov/pmc/articles/PMC5308535/

15. Bi P, Wang J, Hiller JE. Weather: driving force behind the transmission of severe acute respiratory syndrome in China? Intern Med J [Internet]. 2007 [cited 2020 May 4];37(8):550–4. Available from: https://onlinelibrary.wiley.eom/doi/abs/10.1111/j.1445-5994.2007.01358.x

16. Casanova LM, Jeon S, Rutala WA, Weber DJ, Sobsey MD. Effects of Air Temperature and Relative Humidity on Coronavirus Survival on Surfaces. Appl Environ Microbiol [Internet]. 2010 May [cited 2020 May 4];76(9):2712–7. Available from: https://www.ncbi.nlm.nih.gov/pmc/articles/PMC2863430/

17. Chan KH, Peiris JSM, Lam SY, Poon LLM, Yuen KY, Seto WH. The Effects of Temperature and Relative Humidity on the Viability of the SARS Coronavirus [Internet]. Vol. 2011, Advances in Virology. Hindawi; 2011 [cited 2020 May 4], p. e734690. Available from: https://www.hindawi.com/journals/av/2011/734690/

18. Tan J, Mu L, Huang J, Yu S, Chen B, Yin J. An initial investigation of the association between the SARS outbreak and weather: with the view of the environmental temperature and its variation. J Epidemiol Community Health [Internet]. 2005 Mar 1 [cited 2020 May 4];59(3): 186–92. Available from: https://jech.bmj.com/content/59/3/186

19. Doremalen N van, Bushmaker T, Munster VJ. Stability of Middle East respiratory syndrome coronavirus (MERS-CoV) under different environmental conditions. Eurosurveillance [Internet]. 2013 Sep 19 [cited 2020 May 4];18(38):20590. Available from: https://www.eurosurveillance.org/content/10.2807/1560-7917.ES2013.18.38.20590

20. Stensballe LG, Nante E, Jensen IP, Kofoed P-E, Poulsen A, Jensen H, et al. Acute lower respiratory tract infections and respiratory syncytial virus in infants in Guinea-Bissau: a beneficial effect of BCG vaccination for girls: Community based case–control study. Vaccine [Internet]. 2005 Jan 26 [cited 2020 Apr 4];23(10):1251–7. Available from: http://www.sciencedirect.com/science/article/pii/S0264410X04006747

21. de Castro MJ, Pardo-SecoJ, Martinón-Torres F. Nonspecific (Heterologous) Protection of Neonatal BCG Vaccination Against Hospitalization Due to Respiratory Infection and Sepsis. Clin Infect Dis [Internet]. 2015 Jun 1 [cited 2020 May 4];60(11):1611–9. Available from: https://academic.oup.com/cid/article/60/11/1611/356084

22. Niobey FM, Duchiade MP, Vasconcelos AG, de Carvalho ML, Leal M do C, Valente JG. [Risk factors for death caused by pneumonia in children younger than 1 year old in a metropolitan region of southeastern Brazil. A case-control study]. Rev Saude Publica. 1992 Aug;26(4):229–38.

23. KleinnijenhuisJ, Quintín J, Preijers F, Joosten LAB, Ifrim DC, Saeed S, et al. Bacille Calmette-Guerin induces NOD2-dependent nonspecific protection from reinfection via epigenetic reprogramming of monocytes. Proc Natl Acad Sci [Internet]. 2012 Oct 23 [cited 2020 Apr 4];109(43):17537–42. Available from: http://www.pnas.org/cgi/doi/10.1073/pnas.1202870109

24. van den Driessche P, Watmough J. Further Notes on the Basic Reproduction Number. In: Brauer F, van den Driessche P, Wu J, editors. Mathematical Epidemiology [Internet]. Berlin, Heidelberg: Springer; 2008 [cited 2020 May 4], p. 159–78. (Lecture Notes in Mathematics). Available from: https://doi.org/10.1007/978-3-540-78911-6_6

25. James Holland Jones *. Notes On R0. In. Available from: https://web.stanford.edu/~jhj1/teachingdocs/Jones-on-RO.pdf

26. Considerations relating to social distancing measures in response to COVID-19 – second update 23 March 2020 [Internet]. Available from: https://www.ecdc.europa.eu/sites/default/files/documents/covid-19-social-distancing-measuresg-guide-second-update.pdf

27. Yu X, Yang R. COVID-19 transmission through asymptomatic carriers is a challenge to containment. Influenza Other Respir Viruses [Internet], [cited 2020 May 4];n/a(n/a). Available from: https://onlinelibrary.wiley.com/doi/abs/10.1111/irv.12743

28. Privacy Policy. You can manage your preferences in ʻManage Cookies’. Loading web-font TeX/Size2/Regular Skip to main content Thank you for visiting nature.com. You are using a browser version with limited support for CSS. To obtain the best experience, we recommend you use a more up to date browser (or turn off compatibility mode in Internet Explorer). In the meantime, to ensure continued support, we are displaying the site without styles and JavaScript. naturenature medicinebrief communicationsarticle A Nature Research Journal Menu Nature MedicineNature Medicine Search E-alert Submit Login Brief Communication Published: 15 April 2020 Temporal dynamics in viral shedding and transmissibility of COVID-19 Xi He, Eric H. Y. Lau, Peng Wu, Xilong Deng, Jian Wang, Xinxin Hao, Yiu Chung Lau, Jessica Y. Wong, Yujuan Guan, Xinghua Tan, Xiaoneng Mo, Yanqing Chen, Baolin Liao, Weilie Chen, Fengyu Hu, Qing Zhang, Mingqiu Zhong, Yanrong Wu, Lingzhai Zhao, Fuchun Zhang, Benjamin J. Cowling, Fang Li & Gabriel M. Leung. Temporal dynamics in viral shedding and transmissibility of COVID-19 | Nature Medicine [Internet], [cited 2020 May 4], Available from: https://www.nature.com/articles/s41591-020-0869-5

29. Cheng H-Y, Jian S-W, Liu D-P, Ng T-C, Huang W-T, Lin H-H. Contact Tracing Assessment of COVID-19 Transmission Dynamics in Taiwan and Risk at Different Exposure Periods Before and After Symptom Onset. JAMA Intern Med [Internet]. 2020 May 1 [cited 2020 May 4]; Available from: https://jamanetwork.com/journals/jamainternalmedicine/fullarticle/2765641

30. Huaiyu Tian, Yonghong Liu, Yidan Li, Chieh-Hsi Wu, Bin Chen, Moritz U. G. Kraemer, Bingying Li Jun Cai. An investigation of transmission control measures during the first 50 days of the COVID-19 epidemic in China | Science [Internet], [cited 2020 May 4], Available from: https://science.sciencemag.org/content/early/2020/03/30/science.abb6105

31. Signorelli C, Scognamiglio T, Odone A. COVID-19 in Italy: impact of containment measures and prevalence estimates of infection in the general population. Acta Bio-Medica Atenei Parm. 2020 10;91(3-S):175–9.

32. Giordano G, Blanchini F, Bruno R, Colaneri P, Di Filippo A, Di Matteo A, et al. Modelling the COVID-19 epidemic and implementation of population-wide interventions in Italy. Nat Med [Internet]. 2020 Apr 22 [cited 2020 May 4];1–6. Available from: https://www.nature.com/articles/s41591-020-0883-7

33. Tobías A. Evaluation of the lockdowns for the SARS-CoV-2 epidemic in Italy and Spain after one month follow up. Sci Total Environ [Internet]. 2020 Jul 10 [cited 2020 Apr 21];725:138539. Available from: http://www.sciencedirect.com/science/article/pii/S0048969720320520

34. Liu K, Ai S, Song S, Zhu G, Tian F, Li H, et al. Population movement, city closure in Wuhan and geographical expansion of the 2019-nCoV pneumonia infection in China in January 2020. Clin Infect Dis Off Publ Infect Dis Soc Am. 2020 Apr 17;

35. Lasry A. Timing of Community Mitigation and Changes in Reported COVID-19 and Community Mobility — Four U.S. Metropolitan Areas, February 26–April 1, 2020. MMWR Morb Mortal Wkly Rep [Internet]. 2020 [cited 2020 May 4];69. Available from: https://www.cdc.gov/mmwr/volumes/69/wr/mm6915e2.htm

36. Lau H, Khosrawipour V, Kocbach P, Mikolajczyk A, Schubert J, Bania J, et al. The positive impact of lockdown in Wuhan on containing the COVID-19 outbreak in China. J Travel Med. 2020 Mar 17;

37. Prem K, Liu Y, Russell TW, Kucharski AJ, Eggo RM, Davies N, et al. The effect of control strategies to reduce social mixing on outcomes of the COVID-19 epidemic in Wuhan, China: a modelling study. Lancet Public Health [Internet]. 2020 Mar 25 [cited 2020 May 4];0(0). Available from: https://www.thelancet.com/journals/lanpub/article/PIIS2468-2667(20)30073-6/abstract

38. Li C, Romagnani P, von Brunn A, Anders H-J. SARS-CoV-2 and Europe: timing of containment measures for outbreak control. Infection [Internet]. 2020 Apr 9 [cited 2020 May 4]; 1–4. Available from: https://www.ncbi.nlm.nih.gov/pmc/articles/PMC7142270/

39. Quarantine alone or in combination with other public health measures to control COVID-19: a rapid review - Nussbaumer-Streit, B - 2020 | Cochrane Library [Internet], [cited 2020 May 4], Available from: https://www.cochranelibrary.com/cdsr/doi/10.1002/14651858.CD013574/full

40. Sobral MFF, Duarte GB, da Penha Sobral AIG, Marinho MLM, de Souza Melo A. Association between climate variables and global transmission oF SARS-CoV-2. Sci Total Environ [Internet]. 2020 Aug 10 [cited 2020 May 4];729:138997. Available from: http://www.sciencedirect.com/science/article/pii/S0048969720325146

41. Wu Y, Jing W, Liu J, Ma Q, Yuan J, Wang Y, et al. Effects of temperature and humidity on the daily new cases and new deaths of COVID-19 in 166 countries. Sci Total Environ [Internet]. 2020 Aug 10 [cited 2020 May 4];729:139051. Available from: http://www.sciencedirect.com/science/article/pii/S0048969720325687

42. Do weather conditions influence the transmission of the coronavirus (SARS-CoV-2)? [Internet]. CEBM. [cited 2020 May 4], Available from: https://www.cebm.net/covid-19/do-weather-conditions-influence-the-transmission-of-the-coronavirus-sars-cov-2/

43. Wang J, Tang K, Feng K, Lv W. High Temperature and High Humidity Reduce the Transmission of COVID-19 [Internet]. Rochester, NY: Social Science Research Network; 2020 Mar [cited 2020 May 4], Report No.: ID 3551767. Available from: https://papers.ssrn.com/abstract=3551767

44. Liu J, Zhou J, Yao J, Zhang X, Li L, Xu X, et al. Impact of meteorological factors on the COVID-19 transmission: A multi-city study in China. Sci Total Environ [Internet]. 2020 Jul 15 [cited 2020 Apr 21];726:138513. Available from: http://www.sciencedirect.com/science/article/pii/S004896972032026X

45. Şahin M. Impact of weather on COVID-19 pandemic in Turkey. Sci Total Environ [Internet]. 2020 Aug 1 [cited 2020 May 4];728:138810. Available from: http://www.sciencedirect.com/science/article/pii/S0048969720323275

46. Tobías A, Molina T. Is temperature reducing the transmission of COVID-19H? Environ Res [Internet]. 2020 Jul 1 [cited 2020 Apr 27];186:109553. Available from: http://www.sciencedirect.com/science/article/pii/S0013935120304461

47. Sajadi MM, Habibzadeh P, Vintzileos A, Shokouhi S, Miralles-Wilhelm F, Amoroso A. Temperature, Humidity and Latitude Analysis to Predict Potential Spread and Seasonality for COVID-19 [Internet]. Rochester, NY: Social Science Research Network; 2020 Mar [cited 2020 May 4], Report No.: ID 3550308. Available from: https://papers.ssrn.com/abstract=3550308

48. Lund Jensen N, Pedersen HS, Vestergaard M, Mercer SW, Glümer C, Prior A. The impact of socioeconomic status and multimorbidity on mortality: a population-based cohort study. Clin Epidemiol [Internet]. 2017 May 10 [cited 2020 May 4];9:279–89. Available from: https://www.ncbi.nlm.nih.gov/pmc/articles/PMC5436773/

49. Saydah SH, Imperatore G, Beckles GL. Socioeconomic Status and Mortality. Diabetes Care [Internet]. 2013 Jan [cited 2020 May 4];36(1):49–55. Available from: https://www.ncbi.nlm.nih.gov/pmc/articles/PMC3526248/

50. Kim S, Seo YB, Jung E. Prediction of COVID-19 transmission dynamics using a mathematical model considering behavior changes. Epidemiol Health. 2020 Apr 13;e2020026.

51. Oksanen A, Kaakinen M, Latikka R, Savolainen I, Savela N, Koivula A. Regulation and Trust: 3-Month Follow-up Study on COVID-19 Mortality in 25 European Countries. JMIR Public Health Surveill. 2020 24;6(2):e19218.

